# Standardizing and Scaffolding Healthcare AI-Chatbot Evaluation

**DOI:** 10.1101/2024.07.21.24310774

**Authors:** Yining Hua, Winna Xia, David W. Bates, George Luke Hartstein, Hyungjin Tom Kim, Michael Lingzhi Li, Benjamin W. Nelson, Charles Stromeyer, Darlene King, Jina Suh, Li Zhou, John Torous

**Author notes:** Equal contribution, ordered alphabetically (David W. Bates, George Luke Hartstein, Hyungjin Tom Kim, Michael Lingzhi Li, Benjamin W. Nelson, Charles Stromeyer, Darlene King, Jina Suh, Li Zhou). Correspondence*: John Torous, MD, MBI Department of Psychiatry, Beth Israel Deaconess Medical Center, Harvard Medical School, 330 Brookline Ave, Boston, MA, 02446, United States.

## Abstract

The rapid rise of healthcare chatbots, valued at $787.1 million in 2022 and projected to grow at 23.9% annually through 2030, underscores the need for robust evaluation frameworks. Despite their potential, the absence of standardized evaluation criteria and rapid AI advancements complicate assessments. This study addresses these challenges by developing the first comprehensive evaluation framework inspired by health app regulations and integrating insights from diverse stakeholders. Following PRISMA guidelines, we reviewed 11 existing frameworks, refining 271 questions into a structured framework encompassing three priority constructs, 18 second-level constructs, and 60 third-level constructs. Our framework emphasizes safety, privacy, trustworthiness, and usefulness, aligning with recent concerns about AI in healthcare. This adaptable framework aims to serve as the initial step in facilitating the responsible integration of chatbots into healthcare settings.

## Introduction

The rapid rise of chatbots, also known as conversational agents, has garnered substantial interest in the healthcare market. Valued at $787.1 million in 2022, the global healthcare chatbot market is expected to grow at an annual rate of 23.9% from 2023 to 2030.^1^ This expansion is driven by the increasing demand for virtual health assistance, growing collaborations between healthcare providers and industry players, and the acceleration prompted by the COVID-19 pandemic. For example, over 1,000 healthcare organizations worldwide developed COVID-19-specific chatbots using Microsoft’s Healthcare Bot service to manage patient inquiries and reduce the burden on medical staff.^2^ Entering the age of generative artificial intelligence (AI), healthcare chatbots have received even more attention since they enable human-level fluent conversations, have reached physician-level performance on board residency examinations^3^ and comparable performance on other medical examinations and questions^4-,6^ and offer easy ways to train and adapt.

But despite their popularity and potential, evaluating healthcare chatbots poses many challenges.^7-9^ A lack of standardized evaluation approaches has led to diverse and inconsistent methods, making comparing chatbot performance difficult. Rapid technological advancements, particularly in generative AI, outpace existing regulatory frameworks^10^, complicating the establishment of evaluation standards. These new chatbots utilizing generative AI are not constrained by decision trees and are often built on top of larger models, meaning both the output and foundation are not stable. With such a moving target for evaluation, there is no widely accepted guideline or framework for evaluating healthcare chatbots. Developers lack a guide for assessment,^11^ and users often rely on company advertisements or marketing claims.

Several evaluation frameworks^12-22^ have emerged in response to these challenges over the last few years, particularly following the popularity of generative AI. These frameworks vary: some review existing works and regroup metrics into a new structure, others adapt non-healthcare evaluation frameworks for this field, and some focus on narrow sub-directions such as specific specialties or chatbot types. Given the need for a general guiding evaluation framework, a novel approach is necessary. Inspired by a framework^23^ for evaluating health apps, which has now been adopted by the American Psychiatric Association (APA), we crafted a general evaluation framework integrating a literature review and broad stakeholder analyses. This approach involves the perspectives of developers, clinicians, patients, and policymakers to create a comprehensive evaluation structure.

## Methods

As healthcare chatbots face a variety of users, there is no single way to evaluate a chatbot. Factors such as safety and privacy, user preferences, technology literacy, accessibility, and treatment goals are crucial in determining the most suitable evaluation method. In addressing these issues, organizations like the Coalition for Health AI (CHAI) have been working on designing guidelines for trustworthy AI. In April 2023, a group of experts representing diverse stakeholders crafted a blueprint for trustworthy AI implementation guidance.^24^ This blueprint includes seven aspects of trustworthy AI in healthcare: usefulness, safety, accountability and transparency, explainability and interpretability, fairness, security and resilience, and enhanced privacy. But this framework serves more as a theoretical foundation rather than an empirical evaluation framework, and its similarity or overlap with other frameworks remains unclear. Building on the construct definitions in this blueprint and existing evaluation frameworks, we 1) identified a total of 11 evaluation frameworks, 2) extracted all individual questions from these frameworks, 3) removed redundant and non-relevant questions, 4) mapped the remaining questions to CHAI constructs, their subcategories, and constructs not covered by CHAI’s blueprint, 5) improved the evaluation framework structure with stakeholders, including healthcare providers, patients, technology developers, epidemiologists, and policymakers, and 6) further merged and rephrased questions based on assigned constructs.

Due to the absence of a comprehensive review of healthcare chatbot evaluation frameworks, we followed the PRISMA guidelines for selecting and reviewing papers (Appendix A) and gathered 356 questions from the 11 evaluation frameworks (Appendix B). After removing redundant and non-relevant questions (n=35, process detailed in Appendix C), the remaining questions were analyzed for face and construct validity and mapped onto seven priority levels, reflecting the CHAI framework. Subcategories were identified by further clustering questions and reorganizing the framework structure, merging and dividing overlapping questions. This process was modeled as a qualitative factor analysis, where all authors examined and reached a consensus on how the questions were categorized. Based on this refined constructs and framework structure, questions were reanalyzed to form a final list (n=271, listed in Appendix D).

## Results

The final framework (first two levels shown in Figure 1; full framework shown in Appendix E) represents three priority-level constructs, 18 second-level constructs, and 60 third-level constructs. The 271 questions covered 56 third-level constructs. Among these questions, Design and Operational Effectiveness accounted for 108 (40%) questions. Trustworthiness and Usefulness accounted for a similar weight of 107 questions each (39%). The most fundamental level of Safety, Privacy, and Fairness included 56 questions (21%). Subcategories have different levels of granularity, with some categories having only one question and others having many (Appendix F).

**Figure 1:**
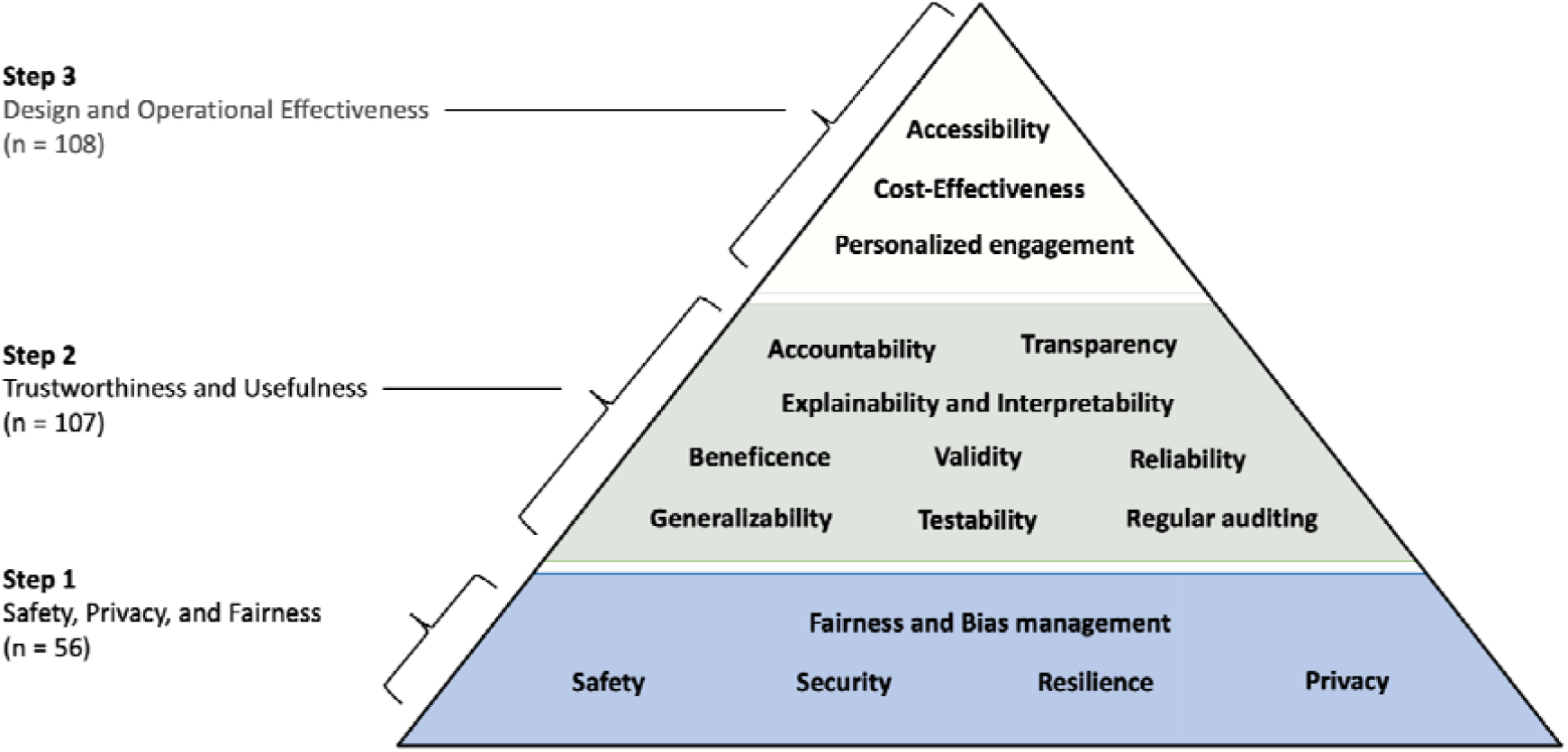
Pyramid for healthcare chatbot evaluation framework. Priority-level constructs are displayed on the left, with second-level constructs within the pyramid.

The rise of generative AI, such as ChatGPT, has expanded interest in healthcare chatbots, placing a pressing need for robust evaluation guidance. Yet the emergence of so many frameworks may create more uncertainty. By assessing the details of numerous frameworks, we were able to simplify and unify different approaches to help inform decision-making. The current framework is designed to be flexible and serve different decision makers around different questions ranging from a designer seeking to create a new chatbot to a patient selecting one from the marketplace. Depending on the user and use case, a different weighting to each construct will be necessary in the same manner that ethical principles offer a scaffold to guide diverse decision making. Our analysis (see Appendix F) suggests that while most frameworks emphasize factors like user experience and task efficiency, stakeholder feedback suggests that a focus on safety and usefulness (see Figure 1) may better match user needs and concerns.

The pyramid structure, similar to Maslow’s Hierarchy of Needs, serves as a visual reminder that evaluation may begin at the base, and progression is likely unnecessary if any level fails to meet the required standards. Still, the user may opt to approach the constructs and questions in any manner that suits their needs.The process of going through these questions will likely facilitate productive dialogue and reveal tensions that must be addressed by the user in order to make the optimal selection. Thus this structure does not itself perform an evaluation but rather serves as a scaffold for evaluation. The same chatbot will be evaluated differently depending on the user and their intent for use, reflecting the flexible nature of this framing. The detailed questions, summarized in Appendix E, are designed to encourage and facilitate dialogue among stakeholders, with responses contextualized within each stakeholder’s unique situation. For instance, some chatbots may collect user conversation histories for training purposes by default. Some patients may find this unacceptable, while others may be comfortable with it. Similarly, developers focused on improving chatbot validity and reliability should not be compelled to conduct user feedback field studies if their research scope explicitly excludes user experience.

## Discussion

Chatbots are increasingly widely used in healthcare, but no comprehensive framework for evaluating their performance has been available. We surveyed the existing frameworks and developed a new framework, using PRISMA guidelines, which we hope will enable future comparisons. This framework is designed to meet the myriad users, use cases, and advances around health AI chatbots by providing a flexible scaffolding to support informed decision making.

Our framework’s foundation in safety, privacy, and fairness is well aligned with recent research raising concerns about these aspects of chatbots. A 2024 review of AI apps concluded these apps may cause harm associated with bias^25^ and the 2023 real-world case of an AI chatbot for eating disorders giving dangerous information to users^26^ highlight the importance of Step 1 (see figure 1) in our framework. Not all AI chatbots are patient facing and the framework is relevant to scaffolding conversations about clinical documentation chatbots, differential diagnosis chatbots, even scheduling chatbots given the core aspects of the framework are relevant. For example, while efforts are underway to identify and address bias in conversational agents,^27^ checking for and identifying bias in any chatbot is a productive first step in considering any conversational agent is a foundational step for avoiding harm.

Likewise, our framework’s second step, trustworthiness, and usefulness, is grounded in recent research. From concerning trends of conversational agents drawing schizophrenia in a stigmatizing manner^28^ to some chatbots providing details on self-harm and how to die by suicide,^29^ it is critical to assess the trustworthiness and usefulness of conversational agents. Given most conversational agents today are trained on social media, not health data,^30^ there is justified concern about the utility of information provided. Additionally, subtle errors can be mixed with correct responses that are difficult for even experts to detect^31^. While there are many approaches to determine trustworthiness and usefulness, and our framework does not dictate which should be employed, the structure ensures a focus on this critical issue.

Our framework also celebrates the success of conversational agents with step three considering factors like their often high degree of accessibility and efforts to personalize content. In placing step three after the prior two, our framework reminds the user to first consider the potential risks and appropriateness of the conversational agent. The majority of frameworks we assessed (see Appendix F) focused on the questions included here in step three. Our approach provides a complimentary means to consider these same questions but in the broader context of steps one and two.

Our framework offers several advantages by synthesizing insights from previous efforts into a new, synergistic model applicable across diverse health conditions and stakeholder groups. Unlike traditional methods that report isolated metrics, our framework reevaluates existing frameworks to distill and integrate them into a comprehensive general guiding framework. It is not designed to challenge or replace any framework and is flexible enough to incorporate new ones that will likely be developed.

A distinctive feature of our framework is its multi-level tree structure, mapping questions into granular constructs without assigning scores to individual questions. This approach facilitates future development of more detailed, domain-specific evaluation methods, using our framework as a reference or guide. Additionally, we aimed to maintain a consistent level of granularity across all levels of the framework, ensuring that each aspect of evaluation is addressed with equal thoroughness.

This approach has several limitations. The framework should be validated prospectively in different contexts to ensure that it is comprehensive and captures important dimensions. There may be additional dimensions that need to be added as the underlying technology quickly evolves, uncovering new issues.

Given the absence of a universal standard for evaluating healthcare chatbots, many parallel review tools have emerged, often failing to capture the full range of important considerations. Our framework addresses this gap, offering a comprehensive, adaptable tool for the evaluation of healthcare chatbots, which we hope will lead to responsible integration of chatbots into healthcare settings. Furthermore, we hope that this review could help guide policymakers to design effective evaluation regulations for healthcare chatbots, both to safeguard the quality of information and provide a clear roadmap for businesses worldwide to further develop tools that improve the quality, efficiency, and effectiveness of care.

This framework presents a starting point that will evolve. Next steps include fully exploring the needs of different users of health AI chatbots and their most common intent/goals. Exploring chatbots beyond the classical medical domains (e.g., nephrology, radiology) and understanding functions across the healthcare ecosystems from scheduling to crisis support will help ensure the framework is responsive to real-world needs. Further work to expand the granularity of individual questions and their focus on users (e.g., developers vs clinicians) will help improve usability. Future endeavors will include a Delphi consensus based on these results in order to engage more stakeholders. Through these efforts, we hope to establish a more rigorous, inclusive, and widely adopted evaluation framework for healthcare chatbots, and enable “apples to apples” comparisons between them.

## Conclusion

This is the first work to develop a structured and adaptable framework for evaluating healthcare AI chatbots, addressing the urgent need for standardized assessment criteria. By synthesizing insights from existing frameworks and diverse stakeholders, we developed a structured approach that prioritizes safety, privacy, trustworthiness, and usefulness. This framework is intended to guide the responsible evaluation and implementation of chatbots in healthcare, helping to ensure their safe and effective use. Future work will focus on validating and refining this framework in different contexts.

## Data Availability

All data produced in the present work are contained in the manuscript

## Declaration of generative AI and AI-assisted technologies in the writing process

During the preparation of this work the author(s) used ChatGPT-4o, web version (accessed 06/26/2024 - 07/02/2024) in order to rephrase some of the framework questions into binary questions (Appendix D). After using this tool/service, the author(s) reviewed and edited the content as needed and take(s) full responsibility for the content of the publication.

## Appendix A: Search Strategy

### A.1. Search Strategy

To identify and evaluate existing frameworks for healthcare conversational agents, we followed the PRISMA guidelines to conduct a systematic review. The literature search was performed across multiple databases to ensure comprehensive coverage of relevant studies. The databases and corresponding search terms were as follows:

| Database | Query |
| --- | --- |
| PubMed<br>(MEDLINE) | ("health"[Title/Abstract] OR "medical"[Title/Abstract] OR "medicine"[Title/Abstract] OR "clinical"[Title/Abstract]) AND ("conversational agent"[Title/Abstract] OR "conversational AI"[Title/Abstract] OR "chatbot"[Title/Abstract] OR "virtual agent"[Title/Abstract] OR "virtual assistant"[Title/Abstract] OR "digital assistant"[Title/Abstract]) AND ("framework"[Title/Abstract] OR "evaluation method"[Title/Abstract] OR "assessment method"[Title/Abstract]) |
| EMBASE | ('health':ti,ab OR 'medical':ti,ab OR 'medicine':ti,ab OR 'clinical':ti,ab) AND ('conversational agent':ti,ab OR 'conversational ai':ti,ab OR 'chatbot':ti,ab OR 'virtual agent':ti,ab OR 'virtual assistant':ti,ab OR 'digital assistant':ti,ab) AND ('framework':ti,ab OR 'evaluation method':ti,ab OR 'assessment method':ti,ab) |
| APA<br>PsychINFO | ("health" OR "medical" OR "medicine" OR "clinical") AND ("conversational agent" OR "conversational AI" OR "chatbot" OR "virtual agent" OR "virtual assistant" OR "digital assistant") AND ("framework" OR "evaluation method" OR "assessment method") |
| The Cochrane<br>Library | ("health" OR "medical" OR "medicine" OR "clinical") AND ("conversational agent" OR "conversational AI" OR "chatbot" OR "virtual agent" OR "virtual assistant" OR "digital assistant") AND ("framework" OR "evaluation method" OR "assessment method") |
| Google<br>Scholar* | ("health" OR "medical" OR "clinical") AND ("conversational agent" OR "conversational AI" OR "chatbot" OR "virtual agent" OR "virtual assistant" OR "digital assistant") AND ("framework" OR "evaluation method" OR "assessment method") |

\*Note: Since Google Scholar does not support advanced search queries, we performed all combinations of searches separately to ensure comprehensive coverage.

### A.2. Inclusion and Exclusion Criteria

The search was restricted to full-length papers published between January 1, 2018, and June 25, 2024. We included studies that developed frameworks for evaluating healthcare conversational agents. We excluded studies introducing new evaluation methods without the intention of providing a structural evaluation framework, such as clinical trials and model development studies.

### A.3. Screening and Selection Process

**Figure A.3.**
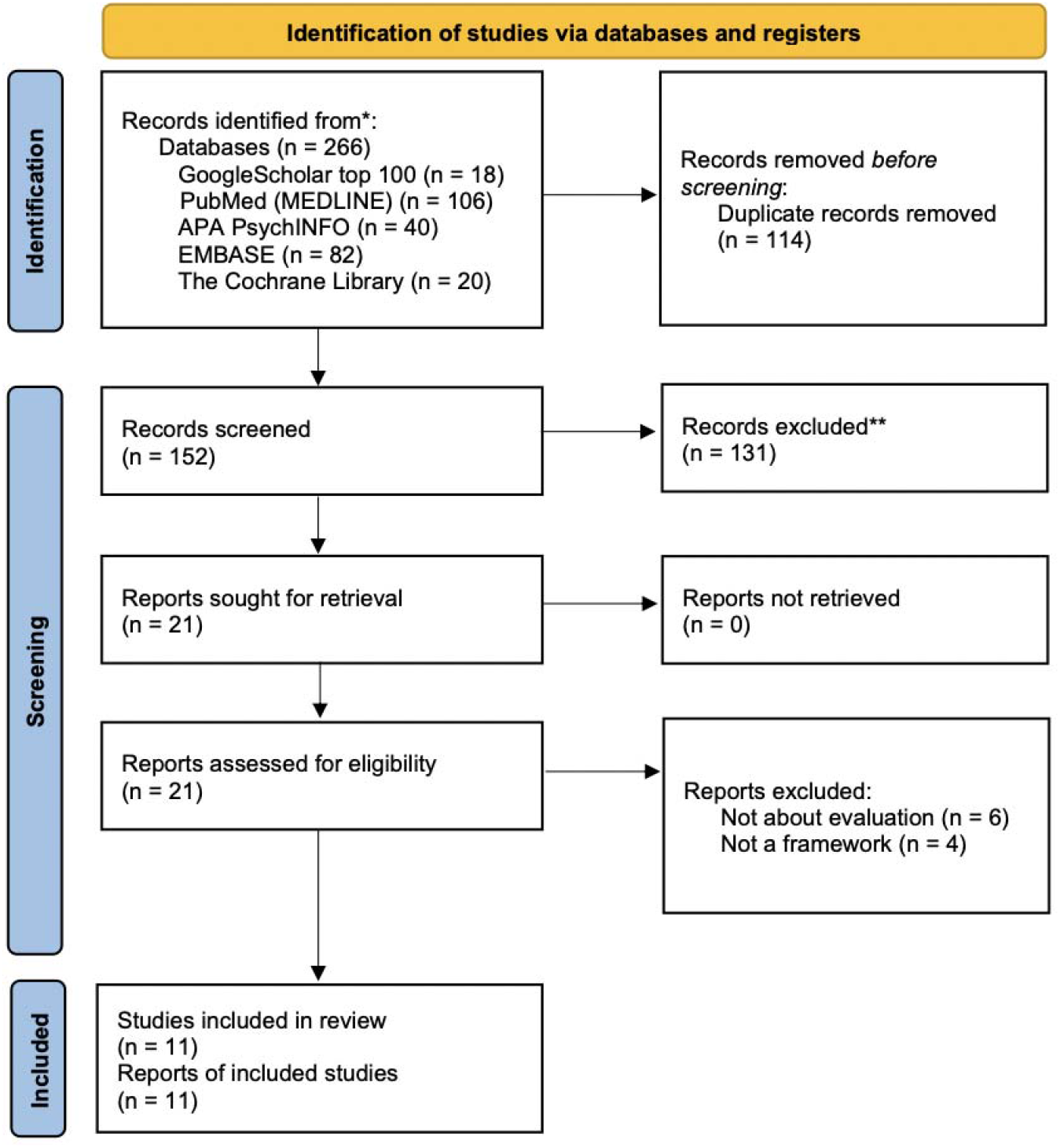
PRISMA Flow Diagram of Study Selection for Evaluation Frameworks of Healthcare Conversational Agents.

The initial search results were screened based on titles and abstracts. Two authors (YH and WX) independently reviewed the titles and abstracts for full-text retrieval, with any discrepancies resolved by discussion with a third reviewer (JT). Full-text articles were then retrieved for further assessment against the inclusion criteria. YH reviewed the full texts and verified them with JT. From the initial 266 records, 152 were screened, and 21 reports were sought for retrieval. After detailed assessment, 11 studies were included in the review, providing a comprehensive evaluation of frameworks for healthcare conversational agents.

## Appendix B: Reviewed Frameworks

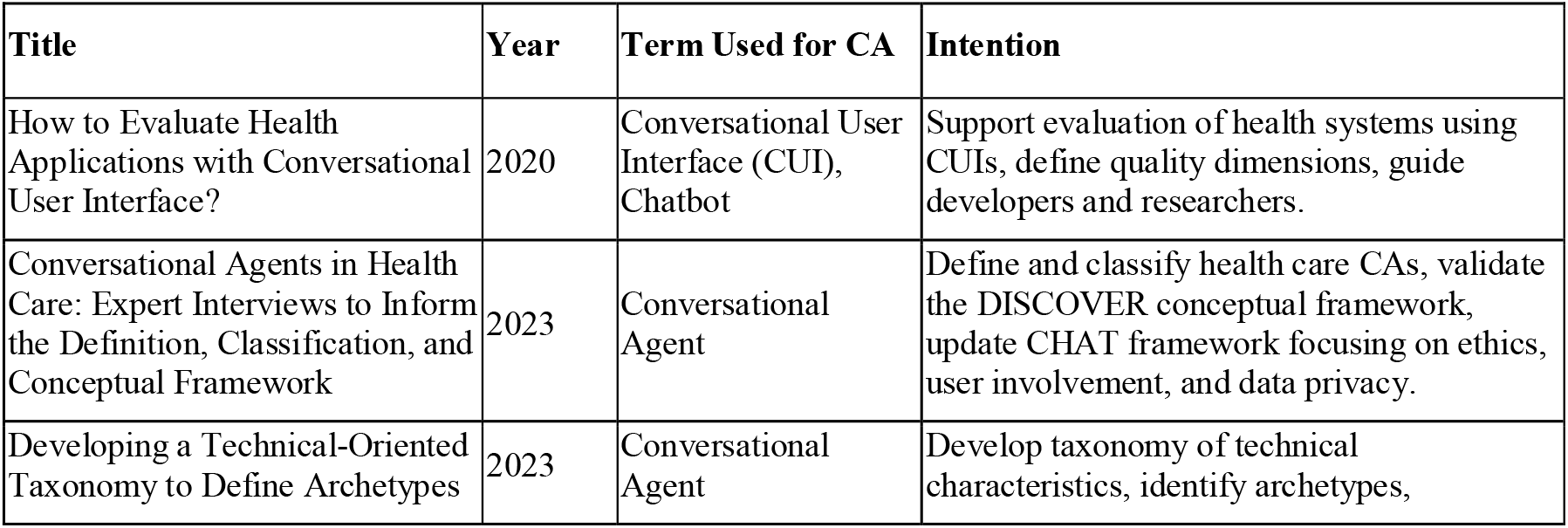
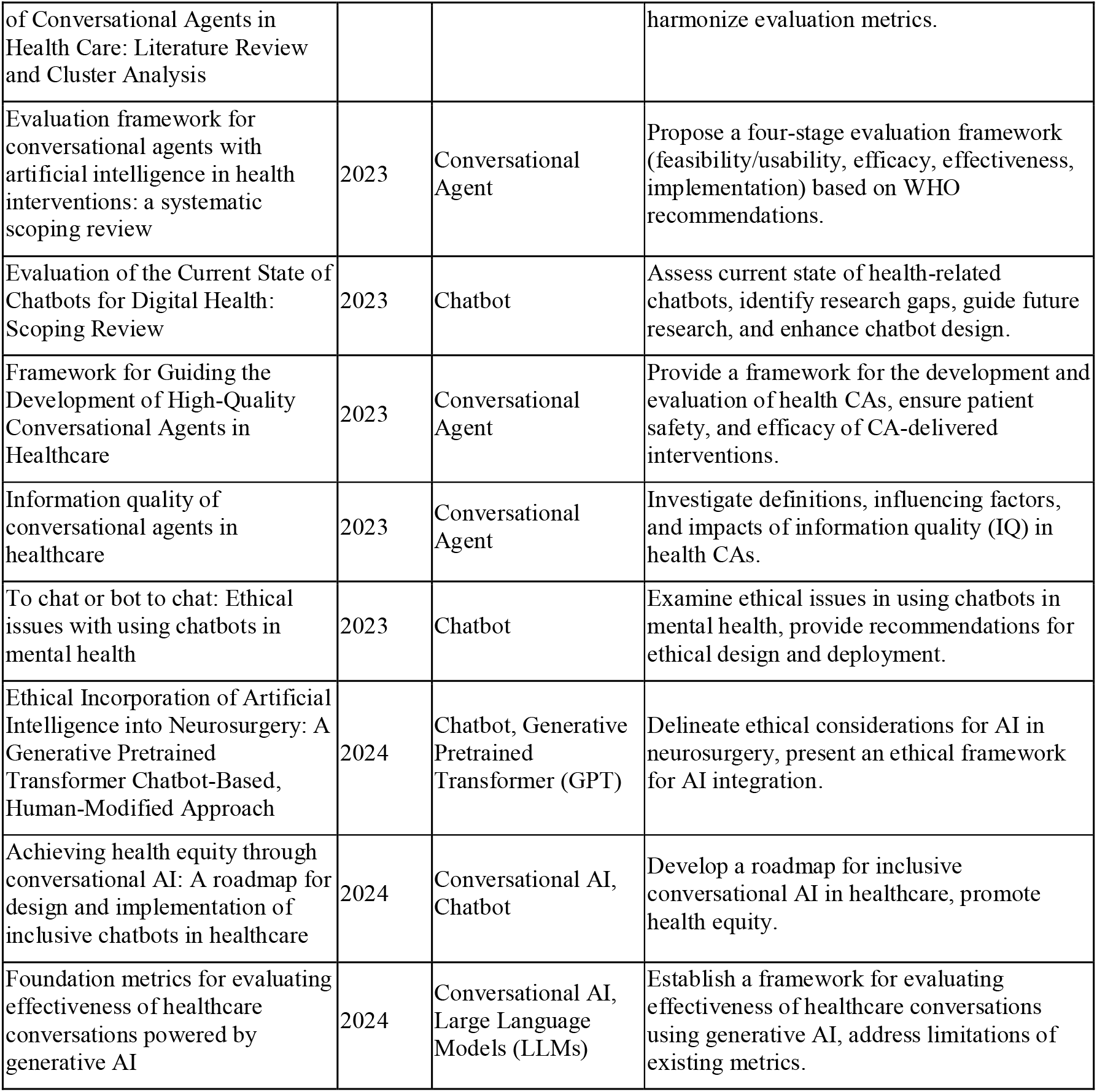

## Appendix C: Details on the review process

We began by summarizing each framework’s intended use to assess specific concepts within a particular domain. The sections detailing the evaluation framework’s questions were then extracted and listed. If the study did not explicitly present evaluation criteria in the form of questions, these criteria were rephrased as questions for clarity. The following steps were taken:

- Describe Use Intention: The purpose and intended application of the framework were articulated, highlighting its relevance and scope.
- Concepts Evaluated: The key concepts and dimensions the framework evaluates were identified and outlined.
- Listing Evaluation Questions: A thorough list of the questions evaluated by the framework was provided. In cases where the study did not present evaluation criteria as questions, these criteria were rephrased into question format for consistency and clarity.

Initially, we broke down questions that contained multiple sub questions. Questions too broad to be constructive were then removed. For instance, we did not include questions such as: “Can strategies or solutions be developed to address problems of CAs?” and “Does the AI system comply with national and international regulations and standards?”

## Appendix D: Extracted Questions and Final Questions

Find in the downloadable supplementary file.

## Appendix E: Tree-structured Framework

1. **Safety, privacy, and fairness**:
  a. **Safety**: prevent worse outcomes for the patient, provider, or health system from occurring as a result of the use of an ML algorithm.
    i. **Outcome proxies appropriateness**: use alternative measures or indicators that accurately reflect the desired health outcomes in the absence of direct measurements.
    ii. **Data provenance**: track and document the origin and history of data, including where it came from and how it has been handled.
      1. **Data Providers**: assign roles and responsibilities to entities like hospital EHRs and patient-generated health data for maintaining safe AI.
      2. **Data Sources**: include various origins of data such as social media and clinical settings.
    iii. **Harm control**: reduce and manage potential risks and negative impacts associated with using a chatbot.
    iv. **Reducing automation bias** (i.e., the tendency to accept automated suggestions without critical evaluation or questioning)
    v. **Critical help**: provide necessary assistance and address negative and help-seeking information.
    vi. **Ethics**: principles and standards that govern the conduct of individuals and organizations, ensuring fairness, privacy, and respect in using ML algorithms in healthcare.
  b. **Security**: maintain confidentiality, integrity, and availability through protection mechanisms that prevent unauthorized access and use
    i. **Protection method**: implement techniques and tools to safeguard data from unauthorized access and threats.
    ii. **Security standard**: follow established guidelines and practices designed to protect data and systems from security breaches.
    iii. **Third-party reliability**: ensure the trustworthiness of external partners or services in maintaining data security and integrity.
  c. **Resilience**: withstand unexpected adverse events or changes in their environment or use
  d. **Privacy**: protect privacy according to standards like HIPAA and GDPR, ensuring user autonomy and dignity.
    i. **Data exchange**: maintain privacy standards for accessing and sharing data with third-party tools, cloud platforms, and other external systems.
    ii. **Data collection and storage:** maintain privacy standards for gathering and securely storing data for future use.
    iii. **Data usage**: maintain privacy standards for using collected data for analysis, decision-making, and improving chatbot algorithms.
    iv. **Privacy Policy**: outline how an organization collects, uses, protects, and shares personal data.
    v. **Data protection**: implement methods to ensure privacy and prevent unauthorized access and breaches.
  e. **Fairness and Bias Management**: ensure the chatbots operate with minimized and acknowledged biases to ensure fair outcomes.
    i. **Systemic Bias**: address biases originating from societal norms and institutional practices.
    ii. **Computational and Statistical Bias**: manage biases arising from the way data is processed and algorithms are designed.
    iii. **Human-cognitive biases**: recognize biases stemming from individual or group perceptions and attitudes.
    iv. **Population bias**: address the issue where certain populations are underrepresented in data, leading to less accurate model performance for those groups.
2. **Trustworthiness and Usefulness**
  a. **Accountability:** ensure those involved in the chatbot’s lifecycle uphold standards of auditability and harm minimization.
  b. **Transparency:** communicate clearly regarding the chatbot’s characteristics and performance throughout its lifecycle.
    i. **Usage Specification**: define how the chatbot should be used.
    ii. **Model Characteristics**: describe the specific features and behaviors of the chatbot.
    iii. **Model Availability**: ensure the chatbot is accessible as needed.
    iv. **Model Limitations**: identify and communicate the boundaries and constraints of the chatbot.
    v. **Data Usage**: explain how data is utilized within the chatbot.
  c. **Explainability and interpretability:**
    i. **Model Explainability**: detail the internal mechanisms and decision-making processes of the chatbot.
    ii. **Model Interpretability**: make the outputs of chatbots clear and meaningful to end-users.
  d. **Beneficence**: ensure chatbot positively impacts its intended outcomes, emphasizing measurable benefits over potential risks.
    i. **Health Outcomes**: focus on improving health results.
    ii. **Clinical Evidence**: use rigorous methods like A/B tests or RCTs to validate effectiveness.
    iii. **User Behaviors**: influence and improve user actions.
    iv. **Intervention**: apply targeted measures to achieve desired outcomes.
    v. **Healthcare System**: integrate effectively within the broader healthcare environment.
  e. **Validity**: ensure the chatbot performs as expected in real-world conditions.
    i. **Data Relevance and Credibility**: use high-quality, pertinent training data.
    ii. **Language Understanding**: ensure the chatbot’s linguistic capabilities are robust.
    iii. **Information Retrieval Accuracy**: accurately retrieve relevant information.
    iv. **Outcome Accuracy**: deliver precise and correct results.
    v. **Task Completion**: effectively complete required functions.
  f. **Reliability**: ensure that the chatbot consistently performs as intended under various conditions and maintains dependable operation over time.
    i. **Failure Prevention**: prevent system failures to maintain functionality.
    ii. **Robustness**: handle unexpected inputs and diverse data without errors.
    iii. **Workflow Integration**: fit seamlessly into existing processes.
    iv. **Reproducibility**: ensure consistent outcomes across different settings.
    v. **Monitoring**: continually check chatbots to ensure proper operation.
    vi. **Up-to-dateness**: keep the system current with the latest information.
  g. **Generalizability**: apply learned patterns to new, unseen data.
    i. **Contextual Adaptability**: function effectively in different environments or clinical contexts.
      1. **Age Group Adaptability**: cater to different age groups.
      2. **Scenario Adaptability**: adapt to various situations.
    ii. **Novel Data Performance**: perform well with new, unseen data.
  h. **Testability**: verify and meet standards for robustness, safety, bias mitigation, fairness, and equity.
    i. **Verifiability**: ensure different attributes can be tested.
      1. **Quantifiability**: measure attributes precisely.
    ii. **Regular Auditing**: measure attributes regularly.
3. **Design and Operational Effectiveness**
  a. **Accessibility**: ensure the chatbot is usable by the intended users, regardless of their abilities, devices, or technical skills, promoting inclusivity and ease of use.
    i. **Versatile access**: provide multiple interaction methods to accommodate user preferences and needs.
      1. **Multi-language**: enable interaction in multiple languages to cater to a diverse user base.
      2. **Different Input and Output Mode**: accommodate various input and output methods, such as text, voice, and visual.
      3. **Multi-platform**: ensure functionality across different platforms, such as web, mobile, and desktop applications.
      4. **Multi-device**: provide compatibility with various devices, including smartphones, tablets, laptops, and desktop computers.
    ii. **User literacy**: ensure the system is usable by individuals with varying levels of technical knowledge and literacy.
    iii. **User experience**: create a pleasant and effective interaction for users.
      1. **Likability**: design the system to be appealing and enjoyable to use.
      2. **Understood by the CA (Conversational Agent)**: ensure clear communication between the user and the chatbot.
      3. **User Engagement**: maintain user interest and active participation.
      4. **Respectfulness**: interact with users in a polite and respectful manner.
      5. **Response Appropriateness**: provide suitable and contextually relevant responses.
      6. **Credibility**: ensure the chatbot’s reliability and trustworthiness.
    iv. **User Interface Design**: create an intuitive and easy-to-use interface for users.
    v. **Simplicity/Ease of Use**: make the system straightforward and user-friendly, minimizing complexity and effort required from users.
  b. **Personalized engagement**: tailor responses based on patient data and preferences.
    i. **Personalization**: customized response based on patient data and preference
    ii. **Anthropomorphism/relationship**: build a human-like relationship with users.
      1. **Relationship Building**: develop a rapport with users.
      2. **Empathy**: show understanding and compassion.
      3. **Humor**: use appropriate humor to engage users.
      4. **Identity**: establish a clear and consistent chatbot persona.
    iii. **User Adherence**: track and analyze how well users follow recommendations, and adjust the chatbot’s strategies based on this data to improve compliance and outcomes
    iv. **Feedback Incorporation**: use user feedback to improve the system.
    v. **Progress awareness**: monitor and respond to the conversation’s context and progress.
      1. **Memory**: support multi-turn or multi-session conversations.
      2. **Strategy Adjustment**: adapt the conversation strategy as needed.
  c. **Cost-effectiveness:** assess whether the chatbot delivers beneficial outcomes at a reasonable cost, providing a better or more economical solution compared to existing methods.
    i. **Comparative Effectiveness**: demonstrate that the chatbot is a better solution than previous methods.
    ii. **Economical Viability**: ensure the system is cost-effective.
    iii. **Environmental Viability**: minimize environmental impact.
    iv. **Task Efficiency**: perform tasks quickly and effectively.
      1. **Appropriate Response Time**: provide timely responses.
      2. **Response Conciseness**: give clear and succinct information.
      3. **Response Relevance**: ensure responses are pertinent to the query.
      4. **Response Practicality**: offer practical and actionable information.
    v. **Workflow Considerations**: integrate smoothly into existing systems.

Questions under constructs such as accessibility assurance and accountability assurance (referenced in Appendix C - Final Questions and Appendix F - Framework Questions Statistics) only assess whether their parent constructs (accessibility and accountability, respectively in this case) are ensured in the evaluation. These placeholder-like subconstructs are not included in this framework for simplicity. Further work is needed to develop questions and future classifications for these constructs, as they are currently overlooked by the literature.

## Appendix F: Framework Question Statistics

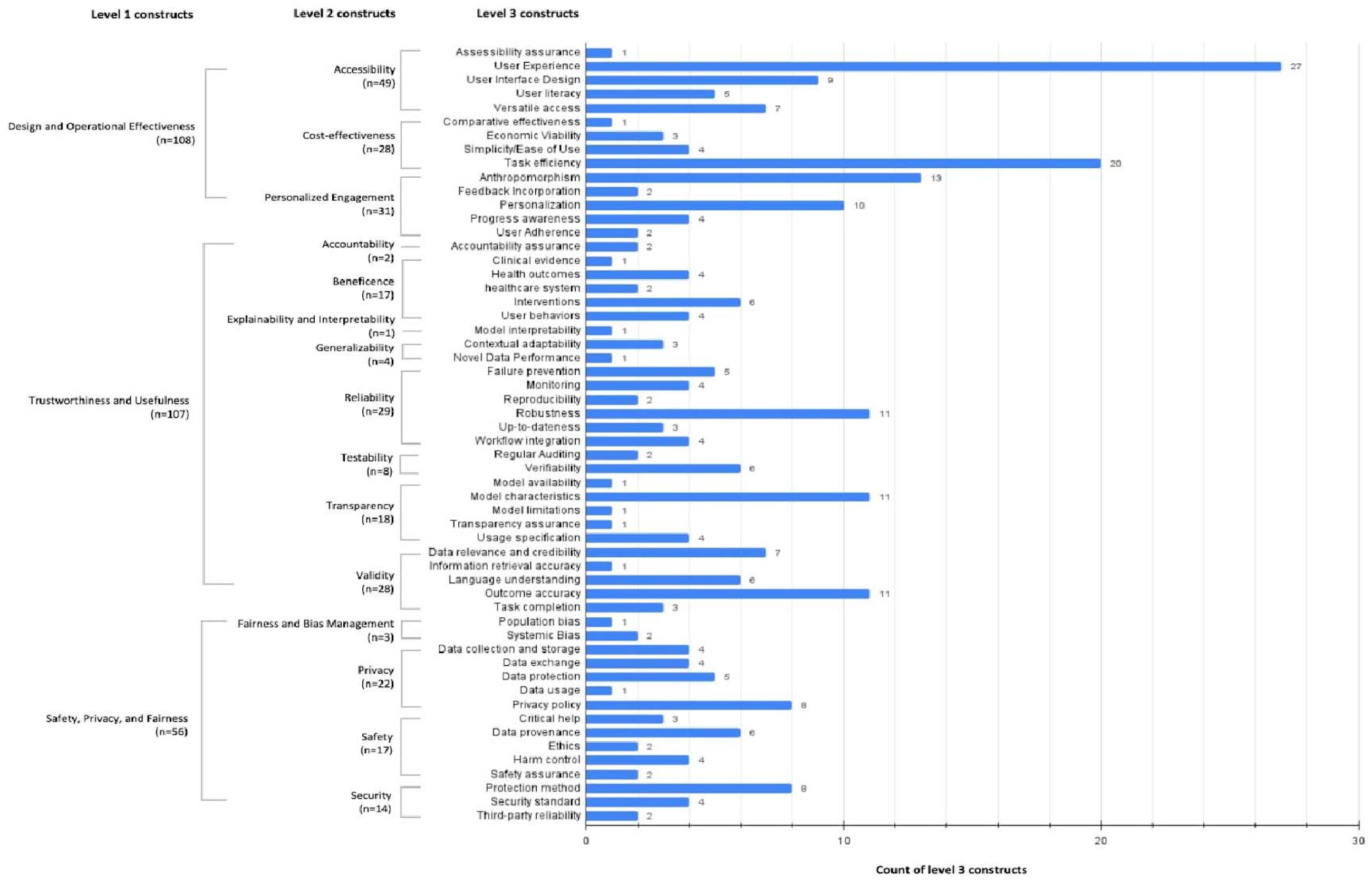

